# Semantic Retrieval of Similar Radiological Images using Vision Transformers

**DOI:** 10.1101/2023.02.16.23286056

**Authors:** Anjali Thakrar, Michael Jayasuriya, Adrian Serapio, Xiao Wu, Eric Davis, Jamie Schroeder, Maya Vella, Jae Ho Sohn

## Abstract

**Background:** Identifying visually and semantically similar radiological images in a database can facilitate the creation of decision support tools, teaching files, and research cohorts. Existing content-based image retrieval tools are often limited to searching by pixel-wise difference or vector distance of model predictions. Vision transformers (ViT) use attention to simultaneously take into account radiological diagnosis and visual appearance.

**Purpose:** We aim to develop a ViT-based image retrieval framework and evaluate the algorithm on NIH Chest Radiographs (CXR) and NLST Chest CTs.

**Materials and Methods:** The model was trained on 112,120 CXR and 111,955 CT images. For CXR, a ViT binary classifier was trained on 4 ground truth labels (Cardiomegaly, Opacity, Emphysema, No Finding) and ensembled to produce multilabel classifications for each CXR. For CT, a regression model was trained to minimize L1 loss on the continuous ground truth labels of patient weight. The ViT image embedding layer was treated as a global image descriptor, using the L2 distance between descriptors as a similarity measure. To qualitatively evaluate the model, five radiologists performed a reader performance study with random query images (25 CT, 25 CXR). For each image, they chose the 5 most similar images from a set of 10 images (the 5 closest and 5 furthest images from the query in model space). Inter-radiologist and radiologist-model agreement statistics were calculated.

**Results:** The CXR model achieved nDCG@5 of 0.73 (p<0.001) and Cardiomegaly mAP@5 of 0.76 (p<0.001) among other results. The CT model achieved nDCG of 16.85 (p<0.001). The model prediction agreed with radiologist consensus on 86% of CXR samples and 79.2% of CT samples. Inter-radiologist Fleiss Kappa of 0.51 and radiologist-consensus-to-model Cohen’s Kappa of 0.65 were observed. A t-SNE of the CT model latent space was generated to validate similar image clustering.

**Conclusion:** Our ViT architecture retrieved visually and semantically similar radiological images.

**Summary Statement:** This study evaluates the efficacy of using ViT based image embeddings for CBIR tasks for CXR and CT images, finding that it performs well on visual and semantic recognition tasks.

**Key Results:**

1. The CXR model achieved nDCG@5 of 0.73 (p<0.001) and Cardiomegaly mAP@5 of 0.76 (p<0.001) among other results for CXR.
2. The CT model achieved nDCG of 16.85 (p<0.001). The model prediction agreed with radiologist consensus on 86% of CXR samples and 79.2% of CT samples.
3. Inter-radiologist Fleiss Kappa of 0.51 and radiologist consensus to model Cohen’s Kappa of 0.65 were observed.

## Introduction

A robust medical content-based image retrieval (CBIR) system provides a platform to identify visually and semantically similar radiological images in a database which could be leveraged to augment educational teaching files, decision support tools, and clinical research cohorts (1). For instance, CBIR systems could be used for educational purposes by removing the need for medical students and residents to analyze an image before composing a text query (2). CBIR systems also hold strong potential for clinical decision support given the prevalence of computer-aided diagnosis in the field of radiology (3). Moreover, content-based methods could be used to retrieve particular types of images to be included in a cohort for a clinical research paper (1).

As the de-facto choice for computer vision-related applications, dominant CBIR techniques use convolutional neural networks (CNNs) to extract image-level descriptors (4). However, inspired by the successes of the transformer architecture in the field of Natural Language Processing, a new attention-based deep learning architecture known as the vision transformer (ViT) was developed whose performance exceeded CNNs on large-scale image recognition tasks (5). In this framework, images are split into fixed-size 2D patches and the resulting sequence of vectors is fed to a standard Transformer encoder (5). The transformer architecture is particularly compelling for radiological applications because it takes spatial relationships between its image patches into account while training (5); we hypothesize that this will allow the model to achieve both a visual and semantic understanding of X-rays and chest CTs that exceeds those found by CNNs.

In this paper, we aim to train a ViT to identify semantically and visually similar images in a database of chest X-Rays (CXRs) and CT scan images and evaluate how the ViT CBIR system compares with radiologists’ perception of similarity with respect to clinical relevance. We hypothesize that the ViT will help bridge the semantic gap in the design of content-based medical image retrieval systems (CBMIR) in the field of radiology.

## Materials and Methods

### Datasets

We consider two large-scale public or limited release datasets: for the X-ray imaging modality, we use the ChestX-ray14 dataset released by the NIH (6) and for the CT-scan imaging modality, we use the National Lung Screening Trial (NLST) dataset (7). We use all 112,120 frontal view X-ray images of the NIH dataset, and all 25,354 images in the NLST CT dataset. We define three X-ray image retrieval tasks, that is for the Cardiomegaly, Opacity, and Emphysema pathologies. The images which contained a pathology among Atelectasis, Effusion, Infiltration, Pneumonia, Consolidation, Edema, Fibrosis, and Pleural Thickening was considered as a positive label for the Opacity image retrieval task. For the Cardiomegaly and Emphysema tasks, we simply consider images that contain the corresponding finding as positive. Any image labeled “No Finding” was considered a negative label for the three tasks. Due to the class imbalance in the dataset, we undersampled the negative examples for each modality to be equal to the number of positive examples. For the CT-scan imaging modality, we chose to define an image retrieval task to distinguish between classes based on the weight (lbs) of each subject. We define three categories to distinguish between: < 150 lbs, between 150 and 200 lbs, and > 200 lbs.

### Data Preprocessing

The dataset for each task is randomly partitioned into 60% training data, 30% validation data for hyperparameter tuning, and 10% test data.

The CXR data was resized to 256 × 256 pixels and center cropped to 224 × 224 pixels, and then pixel normalized by the CheXPert dataset’s mean and standard deviation values. (9). Each image was augmented with a random horizontal flip and random rotation between -5º and 5º (12). Each image was deemed as positive if it contained the target finding for each corresponding image retrieval task (‘Cardiomegaly’, ‘Opacity’, ‘Emphysema’), and negative if the target finding for that image was labeled as ‘No Finding’.

For each CT scan, the middle slice is taken and is windowed according to the corresponding parameters of each DICOM file. The CT images are then subsequently resized to 256 × 256 pixels and are augmented using the same techniques as the CXR data (9). Each scan was placed in the corresponding class dependent on the subject’s weight.

### Model Training

We tune the standard vision transformer described by Dosovitskiy et al which samples patches of size 32×32 from 256×256 images. To construct each dataset’s latent space, we edit the ViT architecture by removing its softmax layer as described in Figure 1. Hyperparameter tuning was done for each task for model selection. The Cardiomegaly vision transformer model was trained for 100 epochs using the SGD optimizer with a learning rate of 6.06e-05 with a batch size of 16 images. Binary cross entropy was the loss function used for model training. The model was trained on a machine with a Linux operating system which has a six-core Intel i7 5930k 3.5-gHz processor (Intel, Santa Clara, Calif), 64 GB of DDR4 SDRAM, and a NVIDIA Pascal Titan X graphical processing unit (Nvidia Corporation, Santa Clara, Calif) with CUDA 8.0 and CuDNN 6.0 (Nvidia). Pytorch (1.11) was used to implement these methods.

**Figure 1:**
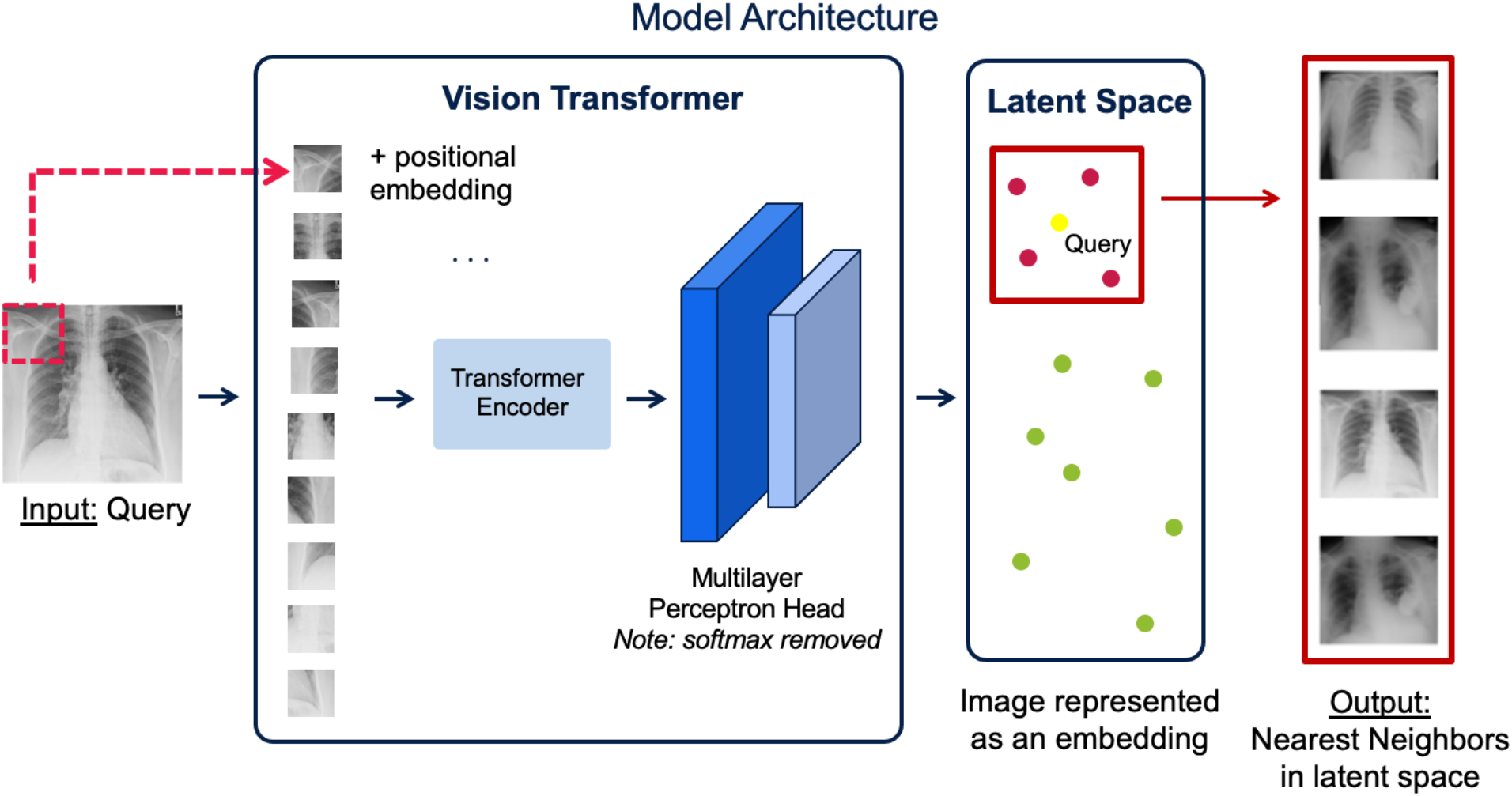
Semantic content-based image retrieval framework. An X-ray/CT-scan image is split into fixed-size patches and fed into a vision transformer to be projected on a latent space. The nearest neighbors are retrieved from the image embedding space.

### Model Evaluation

Each image is passed through the test dataset and the activations of the neurons in the last layer of the network is used as the embedding. For a given query image, we retrieve similar images by computing cosine similarity of each image embedding vector to the query image embedding vector and taking the five smallest. We deemed the retrieved image as relevant if it belongs to the same class as the query image, that is Positive/Negative for the X-ray image retrieval tasks, and the specific weight classes (< 150 lbs, between 150 and 200 lbs, and > 200 lbs) for the CT scan image retrieval task.

For the x-ray imaging modality, we calculate the mean Average Precision (mAP) metric in order to determine whether for each image the relevant images are ranked higher. For the CT images, we use the Normalized Discounted Cumulative Gain (nDCG) score (20), which can be used as a proxy for relevance. In this case, we use the mAP for X-Ray images because it measures discrete, binary relevance, which is useful when using labels as metrics. We use the nDCG for CT images because it allows for the continuous assignment of relevance scores and ranking.

### Reader Performance Study

To evaluate the model’s clinical relevance and ability to retrieve semantically similar images, five board-certified and/or trained resident radiologists (X.W, M.V., E.D., J.S., J.H.S.) with (1, 7, 1, 8, 7) years of post-graduate experience, respectively, participated in a reader performance study. The reader performance study prompted a radiologist to select the 5 most similar images from a group of 10 images, consisting of 5 closest and the 5 furthest from the query in the model space. There were a total of fifty sets of images, composed of 25 sets for both the X-ray and CT-scan imaging modalities. (17, 18, 19)

### Statistical Analysis

Cohen’s Kappa was used to measure the clinical significance of the ViT-based CBIR system against board-certified radiologists and a student’s t-test was used to determine if the mAP scores for each image retrieval task is statistically significant.

## Results

### Dataset

For the X-ray image retrieval tasks, the NIH dataset was composed of 112,120 frontal view X-ray images from 30,805 unique patients. For the Cardiomegaly task, there were a total of 2,193 patients assigned a positive label. For the Opacity class (Atelectasis, Effusion, Infiltration, Pneumonia, Consolidation, Edema, Fibrosis, and Pleural Thickening), there were a total of 15,903 patients assigned a positive label, and for the Emphysema task, there were a total of 1,461 patients assigned a positive label.

For the CT scan image retrieval task, the NLST dataset was composed of 25,354 unique patients. 5,526 of the patients fell under the category of weighing less than 150 lbs. 11,619 patients fell under the category of weighing between 150 and 200 lbs. 8,209 patients fell under the category of weighing greater than 200 lbs. We selected the middle slice of each CT scan taken in the first year that they participated in the NLST study for a total of 111,955 images from 25,354 unique patients.

### Model Evaluation

For the X-Ray imaging modality, the mAP scores for the Cardiomegaly, Opacity, and Emphysema tasks were 0.68 (p < .0001), 0.57 (p < .0001), and 0.787 (p < .0001) respectively. For the CT scan imaging modality, the model achieved an nDCG score of 0.73 (p<0.001). The p-values were calculated by comparing the model’s metric score performance against a model that retrieves images at random with a t-test. Figure 2 illustrates the discriminative capacity of the vision transformer in clustering images of Cardiomegaly for one of the X-ray image retrieval tasks.

**Figure 2:**
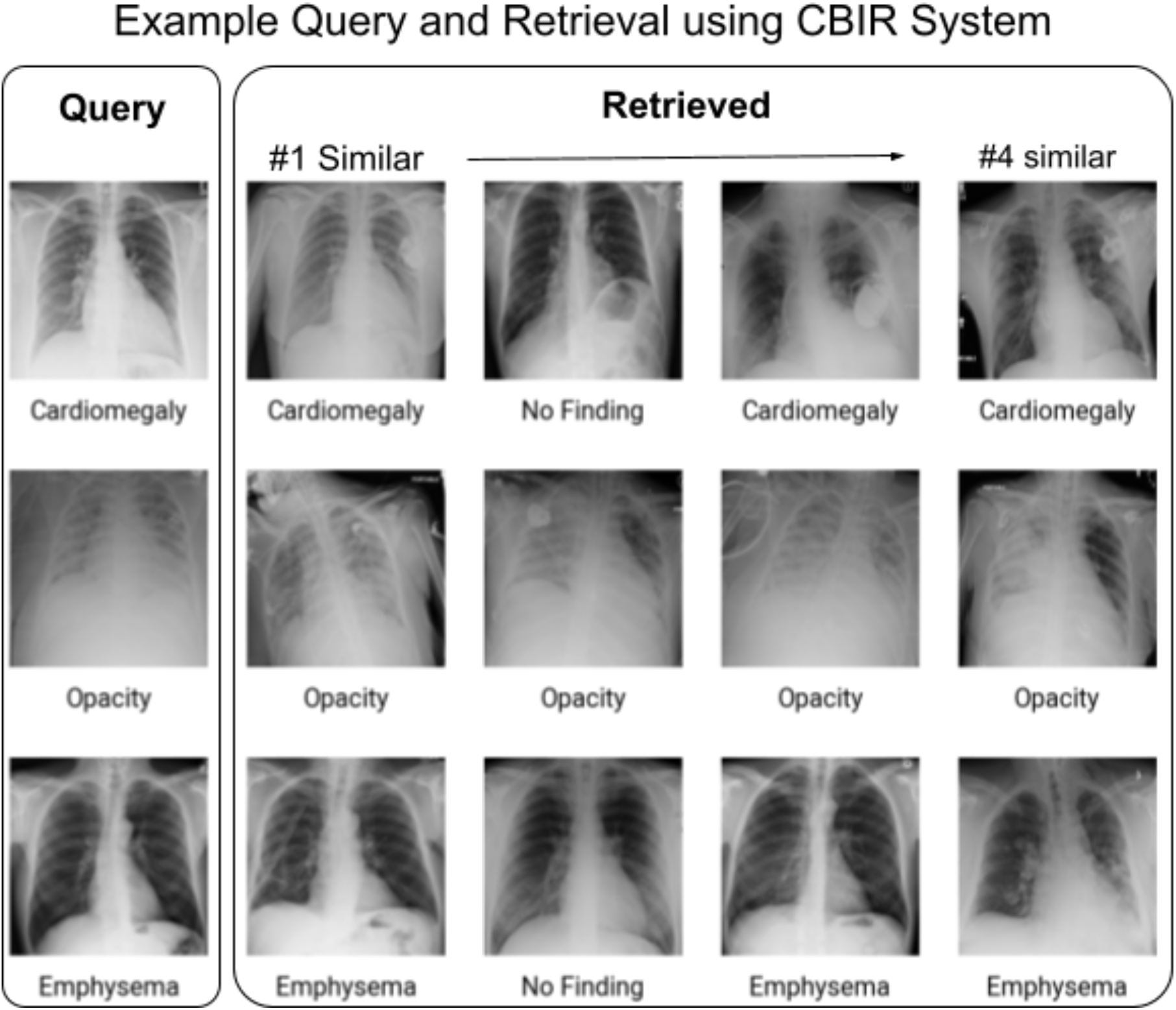
For each task, we use a representative example query and retrieve four images from the ChestX-ray14 dataset using embeddings yielded by the vision transformer. The images in each row satisfy similar diagnostic criteria, with the images in the first row having similar degrees of cardiomegaly, the second row with visually similar opacity, and the images in the third row with similarly severe emphysema. Although the model retrieves two images with mismatching labels, the fact that they are similar to the query image visually demonstrates the ViT’s ability to represent visual relationships between images.

### Reader Performance Study

In the reader performance study, the inter-radiologist Fleiss Kappa of 0.51 was observed, which establishes reliability in the consensus of the radiologists’ classifications. The model prediction agreed with radiologist consensus on 80% of CXR samples and 72.5% of CT samples, and a Cohen’s Kappa of 0.65 was observed between the model and radiologist consensus.

### Error Analysis

The model accuracy on the Cardiomegaly, Opacity, and Emphysema test datasets were 71.4%, 63.4%, 84.52% respectively. In general, the models predict a “No Finding” value more than necessary. For instance, the Cardiomegaly model only correctly classifies about half of the positive cases. The t-SNE in Figure 3 shows qualitatively that there is a group of “No Finding” images that are cleanly separated from the positive “Cardiomegaly” cases, and in the cluster with these positive cases there is pretty significant mixing between the positive and negative class.

**Figure 3:**
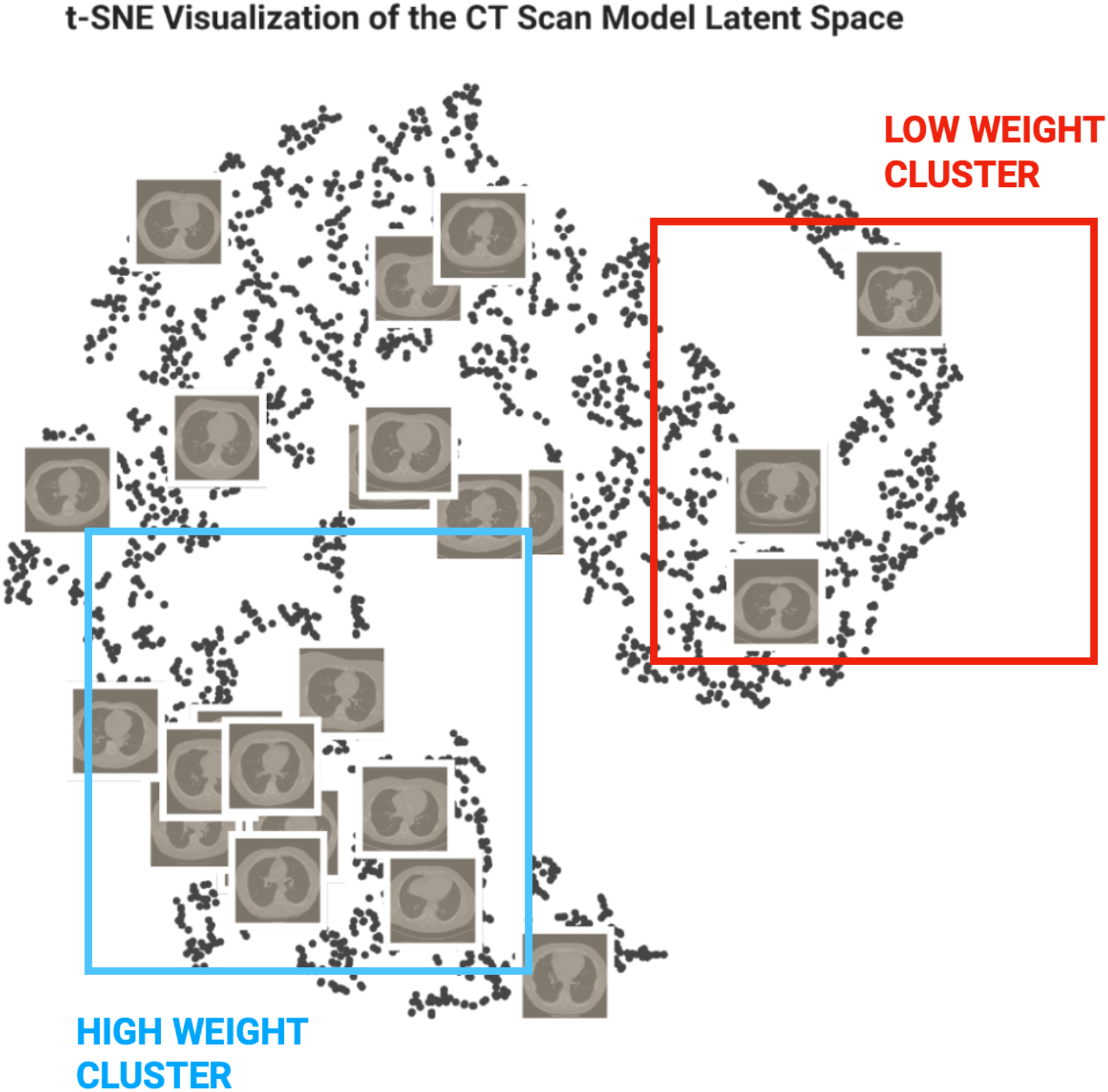
We can visualize the latent space of the model and understand the way the model sees and relates the previously unseen images by graphing our hold-out test set from the NLST dataset. First, we run the images through the trained ViT model to yield embeddings, then dimensionality reduction with them t-distributed stochastic neighbor embedding (t-SNE). The data can be clearly clustered into a high-weight and low-weight region, and the images in these regions not only share similar weight labels but also have similar levels of body fat.

A qualitative analysis of the semantic similarity was performed by a board certified radiologist on selected query images and their closest neighbors in the embedding space by cosine distance. Figure 4 illustrates some example queries and the nearest neighbors on the test split of the ChestX-ray 14 dataset. In the first sample, the clustering was seen to be accurate, with the query image incorrectly labeled as “Cardiomegaly” in the dataset, when it was, in fact, a “No Finding” case. In the second sample, all 5 images are visually similar, with the first and third most similar images being mis-classified as Cardiomegaly. Visually, the lung volume in each of the 5 images is low, indicating that the patients all exhaled. Finally, in the third sample, the model showed a true error case. In the query image, there is a line running on the patient’s right heart border and the patient has a borderline heart size. This may have led the model to falsely understand the image to have a large heart, thus clustering it near cardiomegaly cases.

**Figure 4:**
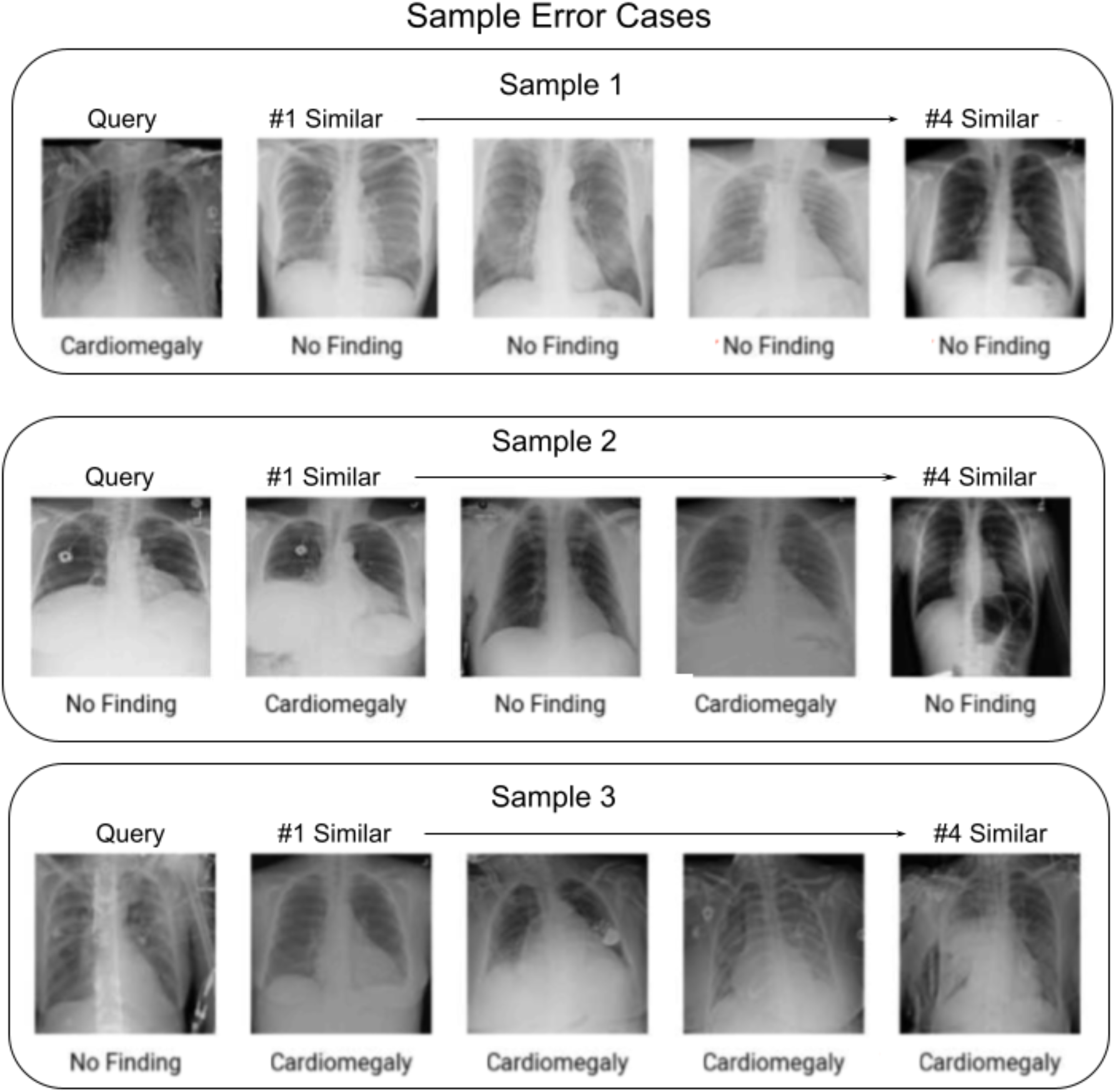
Sample error cases found when generating a set of random queries using the described clustering algorithm. All images are classified using the label it was given in the NIH dataset, and all error analysis stated is subjective and at the discretion of the reviewing radiologist. In sample 1, the query image’s heart is outside of the field of view, so the model may classify the patient as “No Finding” and seems to cluster it accordingly. In sample 2, the #2 and #4 similar images are not visually similar. In sample 3, the query image possibly has an incorrect ground truth label, since the heart is borderline enlarged.

**Table 1.**
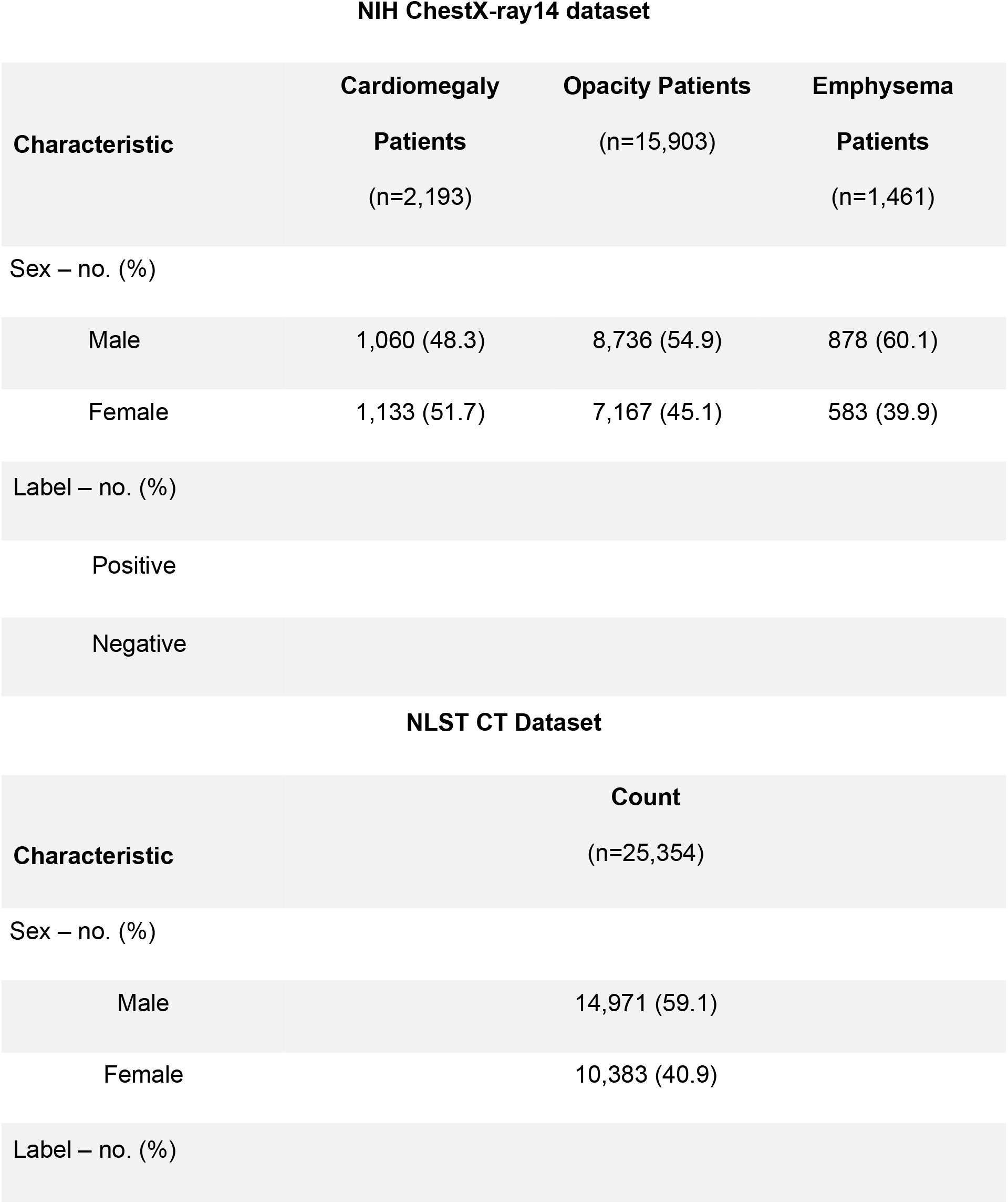
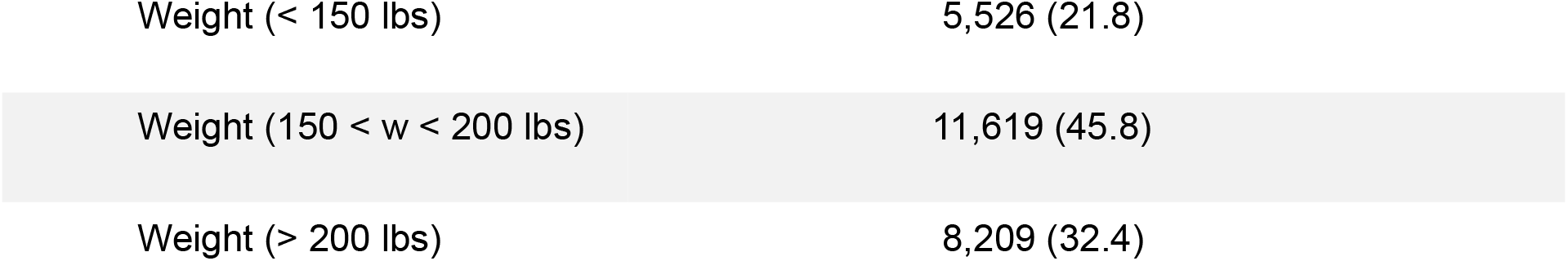
Characteristics of the Datasets. Demographics of each X-ray image retrieval task as well as the CT-scan retrieval task based on patient weight using the ChestX-ray14 and NLST datasets

**Table 2:** Reader performance study metrics and comparison.

|  | Radiologist 1<br>(% agreement) | Radiologist 2<br>(% agreement) | Radiologist 3<br>(% agreement) | Radiologist 4 (%<br>agreement) | Radiologist 5<br>(% agreement) | Average<br>(% agreement) |
| --- | --- | --- | --- | --- | --- | --- |
| CXR | 83.2 | 83.2 | 82.4 | 74.4 | 79.2 | 80.5 |
| CT | 68.8 | 76.8 | 73.6 | 63.2 | 81.6 | 72.8 |

## Discussion

As evidenced by our quantitative results, our study demonstrates metric learning with vision transformers can be used to effectively retrieve X-ray and CT images visually and semantically similar to a query image.

Vision transformers possess the ability to integrate information globally and due to its self-attention modules, attending to image regions that are semantically relevant for classification (5). The biggest promise of ViT models is that they are far more efficient and parallelizable than their CNN counterparts (13, 5). Additionally, while vision transformers lack the inductive biases that make CNN’s train better with less data, they have been shown to learn visual representations that are more robust and generalizable for safety-critical applications such as in the field of healthcare (8). These features make it promising to employ vision transformers for the task of image retrieval (9).

To determine the capacity of our ViT-based CBIR system to query based on visually similar and semantically similar images, we opted to evaluate based on both quantitative and qualitative metrics. For the quantitative aspect, we evaluate the performance of our CBIR system using order-aware metrics (15). We used the mAP in order to determine whether for each image in the test set all of the ground-truth relevant images are ranked higher or not (14).

For the X-ray imaging modality, our model’s statistically significant maP@5 scores on the Cardiomegaly, Opacity, and Emphysema tasks indicate that it is able to, on average, retrieve images that share the same diagnosis as the query image. The model achieved high accuracy for the Cardiomegaly and Emphysema tasks as compared to the Opacity task. This is likely because the opacity task encompasses a wider range of distinct conditions than the other two tasks, which has an overly regularizing effect on the model. We see a similar positive result for the CT scan imaging modality, as the statistically significant nDCG score for the weight image retrieval task indicates that the model consistently retrieves images of similar weights.

Assessing the ability of our system to identify visually similar images also begs the question of what it means for two images to be similar to one another. For the purposes of our experiments, we defined semantic similarity to mean that two images are not only diagnostically similar, i.e. they share the same diagnosis class, but also that they satisfy similar visual criteria for diagnosis. For instance, an image that is borderline Cardiomegaly would be more similar to another image that is borderline Cardiomegaly than an image that has a very pronounced case of Cardiomegaly, even though all these images come from the same class. With visual inspection, we can measure the performance of our model past just classification accuracy.

In practice, this CBIR technology can be used in at least three contexts: diagnostic decision support, education, and research. For example, an attending physician reviewing a case with a resident may want to reference a visually similar case from the past to illustrate a point. Text based tools cannot search with such granularity, but a Vision Transformer can because it searches for differences in image composition, rather than simply using image labels and binary characteristics. It is also easily parallelizable; it does not sacrifice efficiency when searching large databases for the similar image.

CBIR systems can also serve to increase efficiency and accuracy when a radiologist is reviewing an X-ray or CT. By comparing an image to others with visual and semantic similarity, the radiologist is able to rapidly confirm their diagnosis, while also potentially being exposed to other afflictions with similar radiological presentations. When performing a cohort study in radiology, putting together groups of subjects that match one another in presentation can be time consuming. A system with the vision transformer working in the background could easily assemble such cohorts, increasing the velocity of radiology research and allowing more breakthroughs to be made.

There are several limitations noted for our study. For instance, the success of vision transformers are dependent on large amounts of image data (5). However, there exists a paucity of positive image samples that contain abnormal pathologies in the datasets we use, particularly in the ChestX-ray14 dataset released by the NIH (6). While the fine-tuned ViT does show good performance, a model trained on 10M+ radiological images in a self-supervised manner would likely perform even better. As we see from error analysis, the ground truth labels are also often incorrect, especially for the NIH ChestX-ray14 dataset, which likely serves as a ceiling to the performance of our model in this study. Further, the reader performance study was limited by the number of participating radiologists, as well as the small sample size of images reviewed.

A robust, scalable CBIR system based on vision transformers would not only make radiologists much more efficient in clinical decision making and teaching, it could also usher in a new era of evidence based practice. For example, by aggregating similar cases and using other patient metadata such as demographic information, retrospective chart review studies could be computed automatically with the click of the button. The presented method shows that vision transformers have high accuracy in understanding the semantic content of an image, which is particularly powerful in a medical imaging context. More generally, this shows that ViTs are a compelling method to explore in future medical imaging studies. In the future, this study can be extended to provide multi-class classification and clustering to construct a more modular and intelligent system.

Overall, our results demonstrate how this feature of vision transformers allows for the construction of more robust CBIR systems that bridge the semantic gap, the lack of coherence between the information that could be extracted from visual data and a user’s visual perception. The development of robust CBIR systems could facilitate the creation of educational teaching files, decision support tools, and clinical research cohorts, leading to a more integrated approach in human-machine cooperation in the field of radiology (1).

## Data Availability

All data produced are available online

https://nihcc.app.box.com/v/ChestXray-NIHCC

https://cdas.cancer.gov/datasets/nlst/

## Abbreviations

(CBIR): Content-Based Image Retrieval
(ViT): Vision Transformer
(CXR): Chest Radiograph
(CT): Computed Tomography
(mAP): Mean Average Precision
(nDCG): Normalized Discounted Cumulative Gain
(NIH): National Institutes of Health
(NLST): National Lung Screening Trial

## Acknowledgements

The authors thank the Screening Center investigators and staff of the NLST (NLST-297) for providing access to the NLST Chest CT X-Ray dataset. They also thank the NIH Clinical Center for providing public access to the Chest X-Ray 14 dataset.

## Tables

**Supplementary Table 1.**

|  |  |  |
| --- | --- | --- |
| n = 556 | Cardiomegaly | No finding |
| Cardiomegaly | 244 | 33 |
| No finding | 140 | 139 |
| n = 556 | Opacity | No finding |
| Opacity | 244 | 33 |
| No finding | 140 | 139 |
| n = 556 | Emphysema | No finding |
| Emphysema | 244 | 33 |
| No finding | 140 | 139 |

## Figures

**Supplement Figure 1:**
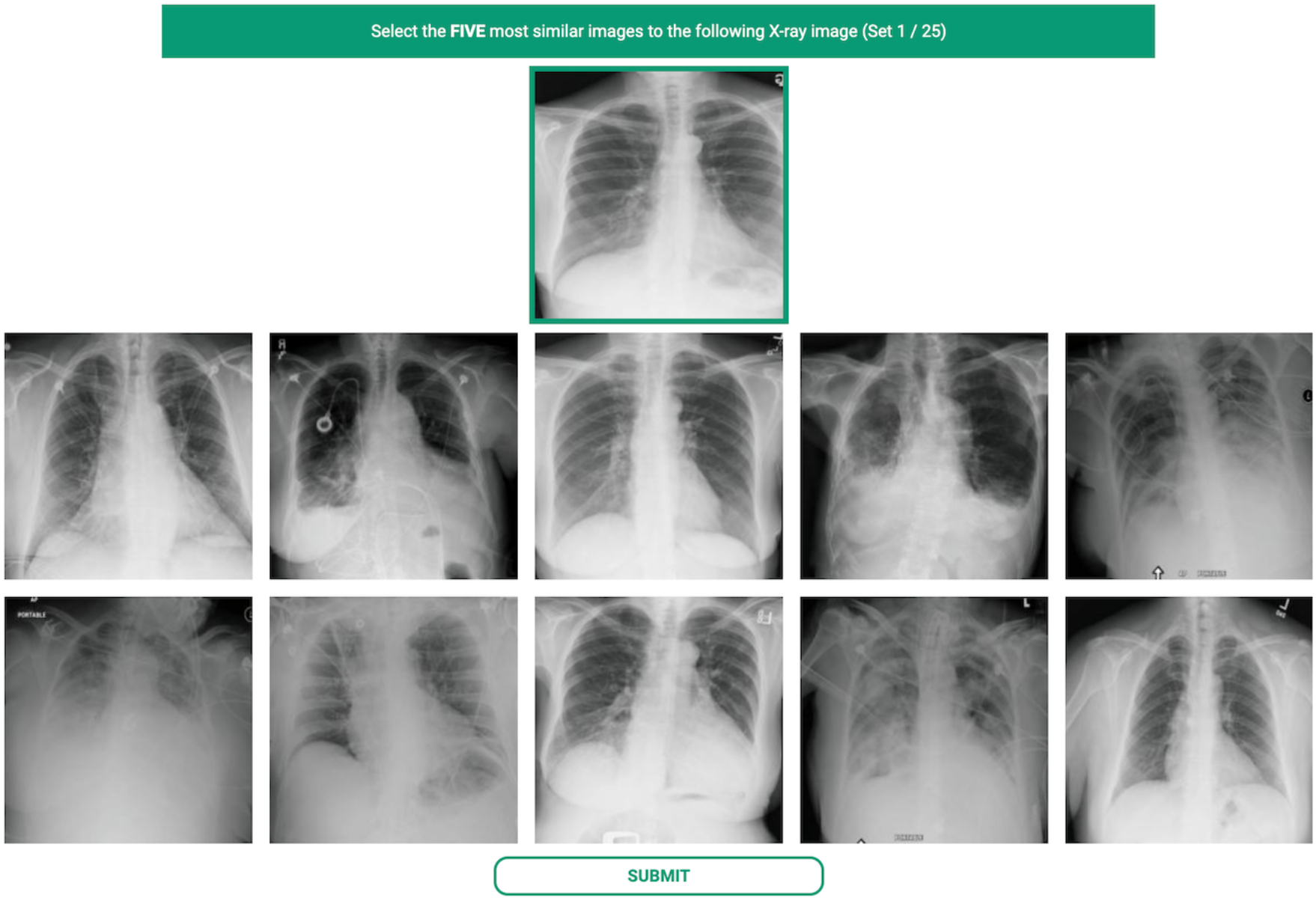
Reader performance study platform used to evaluate the coherence of the model’s retrieval results and radiologist’s perception of similarity based on clinical relevance

**Supplemental Figure 2:**
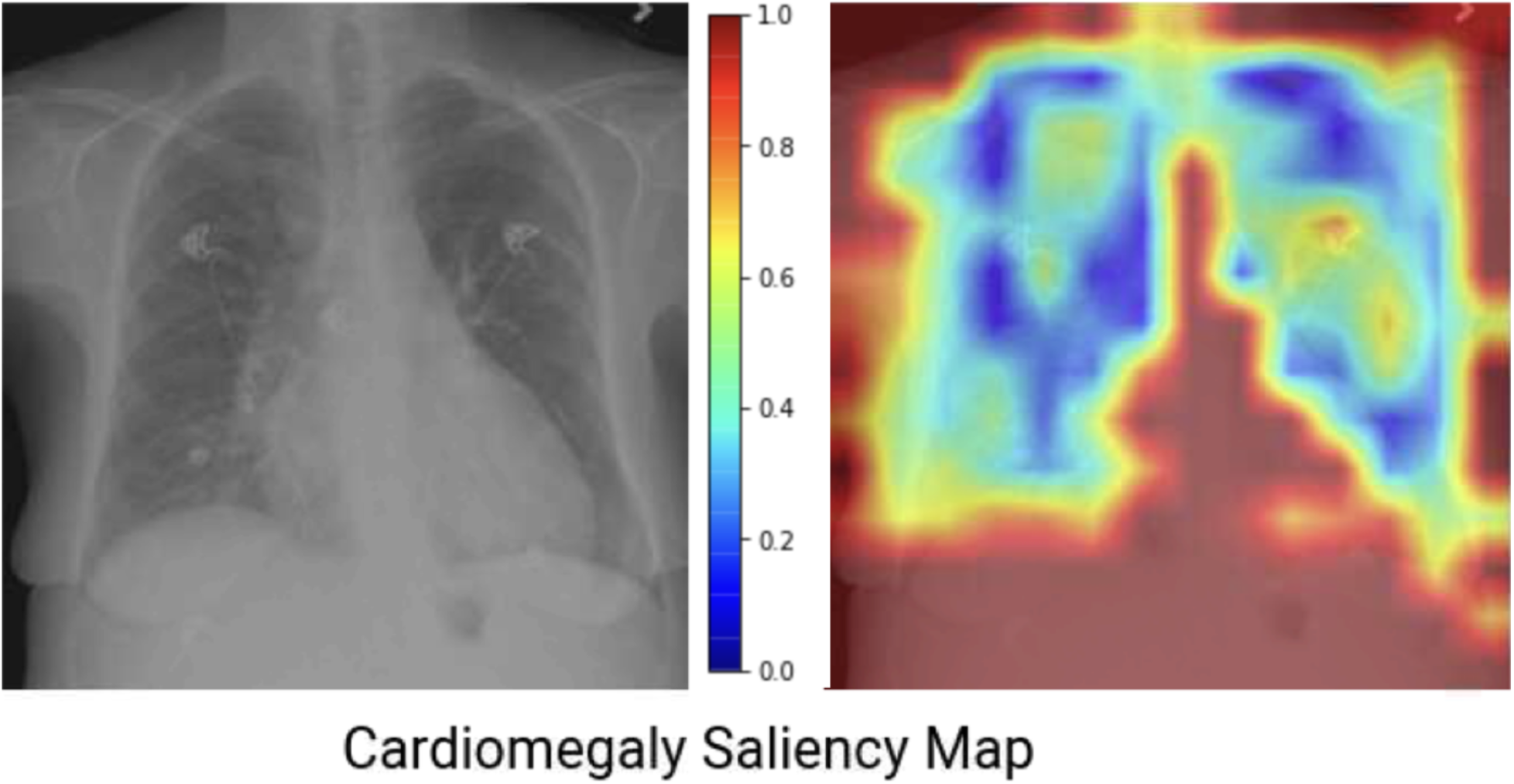
Grad-CAM heatmap is computed using the gradients flowing into the norm1 layer of the ViT. Red indicates regions of high importance to the model’s decision-making. Yellow, green and blue regions are of decreasing importance in the model’s eyes.

## References

1. Müller H, Rosset A, Garcia A, Vallée J-P, Geissbuhler A. Benefits of Content-based Visual Data Access in Radiology. RadioGraphics. 2005;25(3):849–858. doi: 10.1148/rg.253045071.

2. Bucci G, Cagnoni S, De Dominicis R. Integrating content-based retrieval in a medical image reference database. Comput Med Imaging Graph. 1996;20(4):231–241. doi: 10.1016/S0895-6111(96)00016-X.

3. Doi K. Current status and future potential of computer-aided diagnosis in medical imaging. Br J Radiol. 2005;78:S3–S19. doi: 10.1259/bjr/82933343.

4. Wan J, Wang D, Hoi SCH, et al. Deep Learning for Content-Based Image Retrieval: A Comprehensive Study. Proc 22nd ACM Int Conf Multimed. Orlando Florida USA: ACM; 2014. p. 157–166. doi: 10.1145/2647868.2654948.

5. Dosovitskiy A, Beyer L, Kolesnikov A, et al. An Image is Worth 16×16 Words: Transformers for Image Recognition at Scale. arXiv; 2021 Jun. Report No.: arXiv:2010.11929. doi: 10.48550/arXiv.2010.11929.

6. Wang X, Peng Y, Lu L, Lu Z, Bagheri M, Summers RM. ChestX-Ray8: Hospital-Scale Chest X-Ray Database and Benchmarks on Weakly-Supervised Classification and Localization of Common Thorax Diseases. 2017 IEEE Conf Comput Vis Pattern Recognit CVPR. Honolulu, HI: IEEE; 2017. p. 3462–3471. doi: 10.1109/CVPR.2017.369.

7. The National Lung Screening Trial: Overview and Study Design. Radiology. 2011;258(1):243–253. doi: 10.1148/radiol.10091808.

8. Naseer MM, Ranasinghe K, Khan SH, Hayat M, Shahbaz Khan F, Yang M-H. Intriguing Properties of Vision Transformers. In: Ranzato M, Beygelzimer A, Dauphin Y, Liang PS, Vaughan JW, editors. Adv Neural Inf Process Syst. Curran Associates, Inc.; 2021. p. 23296–23308. https://proceedings.neurips.cc/paper/2021/file/c404a5adbf90e09631678b13b05d9d7a-Paper.pdf.

9. El-Nouby A, Neverova N, Laptev I, Jégou H. Training Vision Transformers for Image Retrieval. ArXiv210205644 Cs. 2021; http://arxiv.org/abs/2102.05644. Accessed June 23, 2022.

10. Rajpurkar, Pranav, et al. CheXNet: Radiologist-Level Pneumonia Detection on Chest X-Rays with Deep Learning. arXiv, 25 Dec. 2017, http://arxiv.org/abs/1711.05225.

11. A. Mikołajczyk and M. Grochowski, “Data augmentation for improving deep learning in image classification problem,” in 2018 international interdisciplinary PhD workshop (IIPhDW), 2018, pp. 117–122.

12. Elgendi, M., Nasir, M. U., Tang, Q., Smith, D., Grenier, J.-P., Batte, C., Spieler, B., Leslie, W. D., Menon, C., Fletcher, R. R. et al. (2021), ‘The effectiveness of image augmentation in deep learning networks for detecting covid-19: A geometric transformation perspective’, Frontiers in Medicine

13. Khan, Salman, Muzammal Naseer, Munawar Hayat, Syed Waqas Zamir, Fahad Shahbaz Khan, and Mubarak Shah. “Transformers in vision: A survey.” ACM Computing Surveys (CSUR) (2021).

14. El-Nouby, Alaaeldin, Natalia Neverova, Ivan Laptev, and Hervé Jégou. “Training vision transformers for image retrieval.” arXiv preprint arXiv:2102.05644 (2021).

15. Müller, Henning, Wolfgang Müller, David McG Squire, Stéphane Marchand-Maillet, and Thierry Pun. “Performance evaluation in content-based image retrieval: overview and proposals.” Pattern recognition letters 22, no. 5 (2001): 593–601.

16. Rousseeuw, P. (1987). Silhouettes: a graphical aid to the interpretation and validation of cluster analysis. J. Comput. Appl. Math., 20, 53--65. doi: http://dx.doi.org/10.1016/0377-0427(87)90125-7

17. Faruque, Jessica, Christopher F. Beaulieu, Jarrett Rosenberg, Daniel Rubin, Dorcas Yao, and Sandy Napel. “Content-based image retrieval in radiology: analysis of variability in human perception of similarity.” Journal of Medical Imaging 2, no. 2 (2015): 025501.

18. Wah, Catherine, Grant Van Horn, Steve Branson, Subhransu Maji, Pietro Perona, and Serge Belongie. “Similarity comparisons for interactive fine-grained categorization.” In Proceedings of the IEEE Conference on Computer Vision and Pattern Recognition, pp. 859–866. 2014.

19. Kundel, Harold L., and Marcia Polansky. “Measurement of observer agreement.” Radiology 228, no. 2 (2003): 303–308.

20. Wang, Yining, et al. “A theoretical analysis of NDCG ranking measures.” Proceedings of the 26th annual conference on learning theory (COLT 2013). Vol. 8. 2013.

